# Short Communication: Technological Challenges in Brazilian Healthcare Regulation: Lessons from SUSfácilMG and CORE Saúde MG

**DOI:** 10.64898/2026.10.06.26364925

**Authors:** Lemyson Oliveira Lemos, Letícia A. N. de Sousa, Gleyson José P. Caldeira Silva, Andréa Santos Pinheiro, Janaína Valentim, Tiago de Oliveira Barreto, Felipe Fernandes, Ingridy Barbalho, Nícolas Vinícius Rodrigues Veras, Laura Guerra Pinheiro Reis, João Paulo Q. Santos, Karilany Coutinho, Danilo A. P. Nagem, Jorge Henriques, César Teixeira, Paulo José Carrilho de Sousa Gil, Guilherme Medeiros Machado, Ricardo Valentim

## Abstract

**Background:** Digital systems are central to regulating access to care in Brazil’s Unified Health System (SUS), but fragmented data can hinder coordination.

**Objective:** This study examines the database structure of SUSfácilMG and identifies modernization priorities, using CORE Saúde MG as a comparative reference.

**Methods:** We assessed metadata from 1,586 SUSfácilMG tables, focusing on naming patterns, relationships, and repeated attributes. We then compared architectural features described for CORE Saúde MG.

**Results:** Approximately 40% of the assessed tables repeated descriptive attributes, including patient identifiers, suggesting substantial duplication. Metadata alone cannot establish operational failures, security incidents, or effects on care.

**Conclusion:** A phased review of data models, interfaces, and governance is warranted. Standardized exchange, referential integrity, and controlled access should be evaluated alongside service continuity and data quality.

**Author summary:** Brazil’s public health system relies on digital platforms to organize access to specialized care. In Minas Gerais, SUSfácilMG has supported this process for many years, accumulating adaptations that are reflected in the database structure.

We examined the metadata of 1,586 SUSfácilMG tables, focusing on naming patterns, apparent relationships, and recurring descriptive attributes. Approxim 40% of the tables contained recurring patient-related attributes, indicating struc complexity that may complicate data maintenance and system integration.

Comparison with published descriptions of CORE Saúde MG highlighted opportunities for phased modernization based on shared data structures, interoperability, and governance. Because the analysis was limited to metadata, findings identified structural opportunities rather than evidence of data inconsist security incidents, or effects on patient care.

## Introduction

Regulating access to healthcare is a cornerstone of universality and equity in Brazil’s Unified Health System (Sistema Único de Saúde, SUS), as it translates the constitutional principle of comprehensive care into operational decisions concerning who receives care, where care is provided, and in what order [1]. Access to specialized outpatient care remains a central challenge for SUS managers and researchers, particularly when demand exceeds available capacity and waiting lists operate as mechanisms for prioritizing access [2, 3]. The publication and monitoring of waiting lists have been discussed as strategies for promoting equity and increasing transparency in access to healthcare services [4]. Nevertheless, the literature on healthcare regulation remains strongly centered on normative and institutional dimensions, with comparatively limited attention to how regulatory processes are operationalized and produce care [5].

Healthcare regulation in Brazil operates within a complex institutional and federative framework. The National Regulatory Policy and the broader regulatory framework of the SUS establish mechanisms through which healthcare is coordinated across different levels of complexity, while decision-making depends on articulation and negotiation among the federal, state, and municipal levels of government [6, 7]. Effective regulation, therefore, depends not only on institutional arrangements and service availability but also on the information infrastructure through which regulatory decisions are supported.

From an informational perspective, regulation requires reliable, timely, and comparable data about patients, healthcare facilities, clinical needs, and available capacity, as well as the ability of different systems to exchange these data without loss of meaning. However, health information systems are frequently heterogeneous, institution-specific, and developed for local purposes, resulting in patient information being distributed across multiple systems and healthcare facilities [8]. In Brazil, the persistence of obsolete systems and limited interoperability further contributes to the fragmentation of health information and creates barriers to its integration and effective use [9]. In this context, data governance is essential to ensure that information can be reliably managed, integrated, and used across healthcare systems [10, 11].

These challenges are particularly relevant in long-standing information systems. Healthcare information platforms are often developed incrementally, with modules and functionalities added over time in response to specific institutional and operational needs. This evolutionary process may progressively fragment the architecture and make systems more difficult to maintain and integrate [12]. Incomplete information exchange can also compromise continuity across healthcare settings and generate additional operational and healthcare costs [13]. Thus, the problem extends beyond the existence of regulatory data to how these data are materialized within the structure of long-standing regulatory databases and how such structures may constrain the regulation of access [14].

This issue is particularly relevant in Minas Gerais, where SUSfácilMG has historically supported healthcare regulation across the state. Its long-standing database preserves traces of successive adaptations introduced to accommodate changing regulatory and operational requirements. Examining this structure provides an opportunity to understand how accumulated architectural characteristics may create challenges for system maintenance, data governance, integration, and interoperability. More broadly, legacy systems may accumulate technical debt as successive modifications increase structural complexity and dependencies, making future adaptation progressively more difficult [15–17].

Modernizing such infrastructure requires more than simply replacing existing technologies. Strategies for evolving legacy systems increasingly emphasize modular architectures and the separation of system components, allowing functionalities to be maintained, updated, and integrated more independently [18, 19]. In Minas Gerais, CORE Saúde MG — the Central de Operações para Regulação Estadual (CORE/MG)–developed through a partnership involving the Minas Gerais State Health Department and the Laboratory for Technological Innovation in Health of the Federal University of Rio Grande do Norte (LAIS/UFRN), provides a contemporary architectural reference based on the separation of services and interface-driven components [20]. However, architectural modernisation alone does not demonstrate improvements in waiting times, patient outcomes, or access to care, as interoperability also depends on structural, semantic, organisational, and governance dimensions [21].

Against this background, an important question concerns what the internal structure of a long-standing regulatory database reveals about the challenges involved in modernizing healthcare regulation. Although the literature has extensively discussed healthcare regulation, interoperability, governance, and digital transformation, comparatively less attention has been directed toward the database structures underlying regulatory platforms and the technical legacy that has accumulated throughout their evolution.

This Short Communication examines metadata from 1,586 tables in the SUSfácilMG database to characterize its structural patterns and identify architectural features relevant to maintenance, data governance, and interoperability. Rather than treating database structure as direct evidence of reduced access, patient harm, security incidents, or regulatory non-compliance, the analysis considers metadata as evidence of potential structural challenges associated with the evolution of a long-standing regulatory system [22]. Based on these findings, this study discusses how lessons derived from the SUSfácilMG architecture may inform a phased transition toward more modular and interoperable regulatory infrastructures, using CORE Saúde MG as a contemporary architectural reference.

## Materials and methods

### Study design

This study adopted a descriptive, exploratory design with a qualitative analytical approach, using the metadata of 1,586 SUSfácilMG database tables as the primary material and published literature on CORE Saúde MG as a contemporary architectural reference. The study was conducted in three sequential phases: (i) structural assessment of the SUSfácilMG metadata, (ii) comparison with architectural features described for CORE Saúde MG, and (iii) formulation of priorities for technological modernization.

The analysis focused on structural and architectural characteristics, particularly data organization, service boundaries, interoperability, and security. No clinical or administrative records were examined, and the study did not assess clinical or operational outcomes. Rather, it sought to characterize the accumulated structure of a long-standing regulatory database and discuss its implications for maintenance, integration, and modernization.

### Phase 1: Structural assessment of metadata

Because the source code, complete technical documentation, and patient-level data were not available, the SUSfácilMG database schema was examined through reverse engineering of its metadata. The analysis focused on three structural dimensions:

1. Naming patterns: Table and attribute names were examined to identify recurring naming conventions and prefixes potentially associated with functional groups within the database.
2. Naming patterns: Table and attribute names were examined to identify recurring naming conventions and prefixes potentially associated with functional groups within the database. Apparent relationships: Potential relationships between tables were inferred from key-like attributes, recurring identifiers, and naming conventions. These relationships were treated as provisional because they were derived from metadata patterns rather than verified database constraints.
3. Repeated attributes: Descriptive attributes occurring across multiple tables were identified and quantified according to the number of tables in which they appeared. Examples included patient name, Cadastro de Pessoas Físicas (CPF), and date of birth. This analysis measured attribute recurrence across tables rather than record duplication; therefore, the presence of the same attribute in multiple tables does not imply duplicated rows or patient records. Recurrent descriptive attributes were interpreted as potential maintenance and consistency risks in light of established principles of database normalization and technical debt.

Together, these dimensions were used to characterize the structural organization of the database and identify patterns potentially relevant to its maintenance, integration, and interoperability.

### Phase 2: Comparative Analysis

The structural characteristics identified in SUSfácilMG were compared with the architectural features described in the literature for CORE Saúde MG across four dimensions: data organization, service boundaries, interoperability, and security. Governance was treated as a cross-cutting dimension because it underpins the development and operation of integrated and interoperable health information systems and encompasses the legal frameworks, stewardship mechanisms, data quality standards, and ethical safeguards required for the reliable use of health information.

The comparison was documentary and architectural, based exclusively on the characteristics described in the available literature for CORE Saúde MG. It focused on differences in design rather than system performance; therefore, no benchmarking or assessment of waiting times, clinical outcomes, operational performance, or user experience was conducted. The analysis also distinguished architectural potential from demonstrated performance, recognizing that infrastructural advances alone do not ensure interoperability, which also depends on structural, semantic, organizational, and governance dimensions.

### Phase 3: Formulation of modernization priorities

The findings from the structural assessment and architectural comparison were synthesized to formulate priorities for the modernization of SUSfácilMG. The proposed priorities were organized into four areas: data inventory, normalization, integration, and governance. Their sequencing was informed by approaches to legacy system modernization that emphasize gradual transformation while preserving operational continuity.

A conceptual reference model was also developed to illustrate how shared entities, explicit relationships, and foreign keys can reduce the recurrence of descriptive attributes and support future integration. The model was intentionally abstract and not intended to represent a complete database schema or prescribe the migration of specific SUSfácilMG tables. Integration was positioned after inventory and normalization, reflecting the need to understand and reorganize existing data structures before mapping them to contemporary interoperability approaches.

The resulting priorities were treated as directions for phased modernization rather than an implementation plan. Any future migration would require validation against operational workflows, historical data, technical dependencies, and local constraints prior to implementation.

### Ethical considerations and data protection

The study relied exclusively on structural database metadata. No patient identifiers, clinical records, or other personal data were accessed, extracted, stored, or processed, and no attempt was made to identify individuals or reconstruct clinical histories. Accordingly, the analysis was restricted to the database structure and did not involve the processing of health-related personal data. The requirements of the General Personal Data Protection Law (Lei Geral de Proteçaõ de Dados Pessoais, LGPD) [23] remain applicable to any future access or processing of personal or sensitive health data held in the system [22].

## Results

The metadata assessment identified recurring descriptive attributes across multiple SUSfácilMG tables, particularly patient- and location-related fields. These patterns indicate that similar descriptive information is represented in parallel database structures, suggesting opportunities for consolidating shared entities. From a database design perspective, such recurrence may increase maintenance complexity and the potential for inconsistent values when the same information must be updated across multiple structures [17]. However, because individual records were not examined, the analysis does not demonstrate that data inconsistencies occur in practice.

The organization and number of tables also revealed substantial structural complexity, including naming patterns and inferred relationships that may require careful mapping when integrating SUSfácilMG with external health information systems. These findings reflect the characteristics of the database schema only; metadata alone cannot establish whether specific tables are active or obsolete, how dependencies are implemented at the application level, or whether the system follows a particular software architecture.

Recurring fields potentially associated with sensitive information were identified across schemas. Their presence indicates database locations that may warrant consideration in future assessments of access control, retention auditing and data governance. This study did not evaluate these controls, identify security incidents, or assess compliance with the General Personal Data Protection Law (Lei Geral de Proteção de Dados Pessoais, LGPD) [22, 23].

Figure 1 illustrates the structural pattern identified in SUSfácilMG alongside a conceptually normalized reference model. In the proposed representation, shared entities are connected through keys, reducing the recurrence of descriptive attributes and simplifying their structural organization. The model is conceptual and does not represent a complete database schema or prescribe the migration of individual tables.

**Fig 1.**
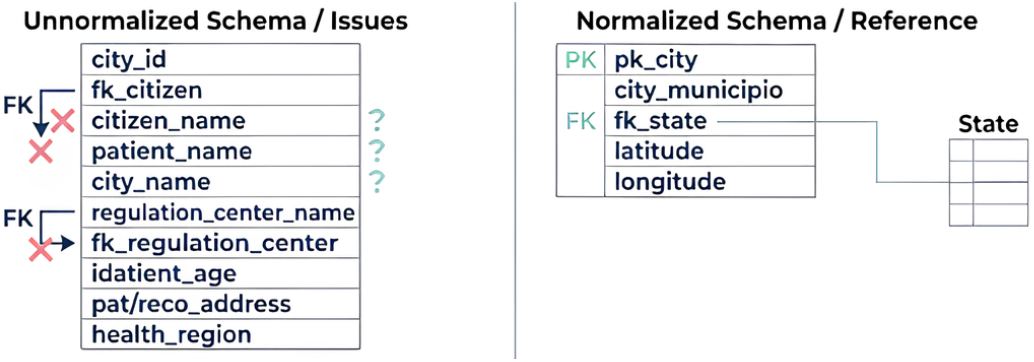
Comparison between recurring descriptive attributes identified in the SUSfácilMG database structure and a conceptual normalized reference model proposed for modernization. **Source:** Author’s own elaboration (2026).

## Discussion

### The CORE Saúde MG model as a reference for digital governance

Published descriptions of CORE Saúde MG provide a contemporary reference for interpreting the structural findings observed in the SUSfácilMG. CORE Saúde MG is a multi-region solution based on service separation and system integration, with functionalities supporting interoperability, real-time monitoring, queue management, transparency, and process auditability [24]. Such service boundaries may facilitate independent component evolution when interfaces and data responsibilities are clearly defined, consistent with interface-driven approaches to legacy system modernization [20].

In this context, the recurring attributes and structural complexity identified in SUSfácilMG highlight areas in which clearer data ownership, shared identifiers, and well-defined interfaces may support future modernization. Published evidence indicates that interoperable platforms and integrated information can strengthen monitoring and decision-making [10], and emerging approaches, including machine learning, have also been investigated in Brazilian healthcare regulation [25].

These architectural possibilities depend on effective governance of data. Repeated patient attributes raise questions regarding authoritative records, access control, corrections, and auditability. However, the present findings do not demonstrate security failures or non-compliance with the LGPD. More broadly, technical infrastructure alone does not ensure interoperability, which also depends on semantic, organizational, and governance arrangements [10, 11, 21].

Accordingly, CORE Saúde MG should be interpreted as an architectural reference rather than a performance benchmark. The available evidence supports the comparison of design characteristics but not conclusions regarding superior operational or clinical performance. Such conclusions would require direct evaluation using indicators such as data quality, request-processing time, system availability, and equity of access.

### Strategic guidelines for the modernization of health regulation

The findings support a phased approach to modernization, beginning with an inventory of tables, data flows, interfaces, and actual system use. This initial assessment could help identify authoritative records for core entities, clarify dependencies, and guide subsequent normalization and integration efforts. Incremental migration, supported by validation and rollback mechanisms, may reduce the risks to service continuity during legacy system transformation [18, 19].

Interoperability should follow a detailed assessment of existing data structures and semantics. Standards, such as HL7 FHIR, may provide a framework for selected information exchanges, but their adoption requires local identifiers, fields, and terminologies to be mapped and validated against connected systems [26]. This is particularly relevant when legacy structures do not map directly to contemporary interoperability models [15, 16].

Modernization should also incorporate data governance and security throughout the transition, including the definition of data ownership, access controls, authentication, auditability, retention, and incident response procedures. When automated prioritization is considered, its implementation would require validation using appropriate local data and continued monitoring for errors and potential inequities.

These priorities should be understood as strategic directions derived from structural analysis rather than a prescriptive implementation roadmap. Their feasibility and impact must be evaluated against the system’s actual workflows, technical dependencies, operational requirements, and local governance arrangements.

## Conclusion

The assessment of 1,586 SUSfácilMG tables identified substantial recurrence of descriptive attributes, highlighting opportunities for structural review, consolidation of shared entities, and improved interoperability. These findings support the need for a targeted technical assessment but do not establish the effects on patient safety, waiting times, system performance, or legal compliance.

CORE Saúde MG provides a contemporary architectural reference for considering more modular and interoperable approaches to health regulation. Based on the structural findings, modernization could prioritize data-model review, governance of shared identifiers, documented interfaces, and privacy and security controls. Standards, such as HL7 FHIR, may support selected information exchanges when accompanied by agreed semantics and data models adapted to the requirements of the SUS.

Because the analysis was limited to metadata and published architectural descriptions, further investigation is required to validate the actual database use, technical dependencies, and migration feasibility. Future studies should also assess whether the implemented changes improve data quality, service continuity, regulatory processes, and equitable access in practice.

## Data Availability

The data underlying the findings of this study consist of structural metadata from the SUSfàcilMG database, including table names, attribute names, and naming conventions. No patient-level, clinical, or other personal data were accessed or analysed. The metadata were provided for the purposes of this study and are not publicly available because they relate to the internal structure of a governmental health information system. Requests for access should be directed to the Secretaria de Estado de Saàde de Minas Gerais (SES-MG), subject to institutional authorization and applicable data governance requirements.

## Acknowledgments

We sincerely thank the Laboratory for Technological Innovation in Health (LAIS) at the Federal University of Rio Grande do Norte (UFRN), the State Department of Health of Minas Gerais, Brazil, and the Ministry of Health of Brazil for their support of this study. The authors express their gratitude to Bruna Fernandes de Araújo for carefully translating the original text into English.

